# Lung function from school age to adulthood in primary ciliary dyskinesia

**DOI:** 10.1101/2021.07.08.21260172

**Authors:** Florian S Halbeisen, Myrofora Goutaki, Eva S L Pedersen, Ben D Spycher, Israel Amirav, Mieke Boon, Cohen-Cymberknoh Malena, Suzanne Crowley, Nagehan Emiralioglu, Eric G Haarman, Bulent Karadag, Cordula Koerner-Rettberg, Philipp Latzin, Michael R. Loebinger, Jane S Lucas, Henryk Mazurek, Lucy Morgan, June Marthin, Petr Pohunek, Francesca Santamaria, Nicolaus Schwerk, Guillaume Thouvenin, Panayiotis Yiallouros, Kim G Nielsen, Claudia E Kuehni

## Abstract

Primary ciliary dyskinesia (PCD) presents with symptoms early in life and the disease course may be progressive, but longitudinal data on lung function are scarce. This multinational cohort study describes lung function trajectories in children, adolescents, and young adults with PCD. We analysed data from 486 patients with repeated lung function measurements obtained between the age of 6 and 24 years from the international PCD Cohort (iPCD) and calculated z-scores for forced expiratory volume in the first second (FEV_1_) and forced vital capacity (FVC) using the Global Lung Function Initiative 2012 references. We described baseline lung function and change of lung function over time and described their associations with possible determinants in mixed-effects linear regression models. Overall, FEV_1_ and FVC z-scores declined over time (average crude annual FEV_1_ decline was -0.07 z-scores) but not at the same rate for all patients. FEV_1_ z-scores improved over time in 21% of patients, remained stable in 40% and declined in 39%. Low BMI was associated with poor baseline lung function and with further decline. Results differed by country and ultrastructural defect, but we found no evidence of differences by sex, calendar year of diagnosis, age at diagnosis, diagnostic certainty, or laterality defect. Our study shows that on average lung function in PCD declines throughout the entire period of lung growth, from childhood to young adult age, even among patients treated in specialized centres. It is essential to develop strategies to reverse this tendency and improve prognosis.

**Take home message:** Lung function in children with PCD is reduced by age 6 years and further declines during the growth period. It is essential to develop strategies to improve prognosis.

## Introduction

Primary ciliary dyskinesia (PCD) commonly leads to chronic upper and lower airway disease due to impaired mucociliary clearance. Clinical course is variable, but many patients present with neonatal respiratory distress, develop bronchiectasis already in preschool age, and some progress to end stage lung disease with respiratory failure and transplantation in adulthood.[1-6] In a systematic review covering the period 1980 to 2017, we identified 24 small studies on lung function in patients with PCD.[7] Twelve studies had longitudinal lung function data, mostly from adult patients. Among these, four studies reported that lung function remained stable over time and four concluded that it deteriorated. The largest study reported that FEV1 improved over time in 10% of Danish patients, remained stable in 57%, and declined in 34%.[8] Models of lung function changes over a lifetime distinguish between lung function at birth, lung growth during childhood, a short plateau phase and the long period of lung function decline.[9, 10] The pattern of lung function growth in early years determines lung function later in life, and also life expectancy.[11, 12] Combining patients of different ages in one model does not allow to disentangle these phases and identify possible risk factors. For people with PCD, only two studies focused on lung function growth during childhood.[13, 14] One, with 137 children from specialised PCD clinics in North America, found a variable time course with ultrastructural defects highlighted as the main determinant of heterogeneity.[13] The second, with 158 children from three countries, reported variable lung function trajectories during childhood but no difference between centres.[14]

We have previously presented cross-sectional lung function data from the international PCD cohort study (iPCD) and showed that forced expiratory volume in the first second (FEV1) and forced vital capacity (FVC) were reduced compared to Global Lung Function Initiative 2012 (GLI) reference values in patients of all ages.[15] The design of that study did not permit investigation of lung function changes over time, nor did it allow to identify factors that influence lung function trajectories.

The present study describes lung function trajectories in an international cohort of children and young adults with PCD from age 6 to 24 years. We compared FEV1 and FVC of study participants with the GLI reference values at baseline and investigated changes in lung function over time, i.e. whether z-scores improved, remained stable, or decreased. Second, we investigated a range of potential determinants for their association with lung function at baseline or with subsequent change of lung function over time.

## Methods

### Study design and study population

The iPCD Cohort is a large international dataset of patients with PCD set up during the EU FP7 project “Better Experimental Screening and Treatment for Primary Ciliary Dyskinesia” (BESTCILIA) and expanded during the COST-Action BEAT-PCD.[16, 17] The current analysis includes data from patients aged 6 to 24 years with multiple measurements on FEV_1_ and FVC that were delivered, cleaned, and standardized by March 2019. Data came from routine follow-up in PCD clinics and were extracted from hospital records or exported from regional or national PCD registries.[17] When necessary, principal investigators obtained ethics approval and informed consent or assent in their countries to contribute observational pseudonymised data.

### PCD diagnosis

PCD diagnosis remains challenging and has evolved over the years.[18] Guidelines recommend a combination of tests,[19] but test availability differs between countries and centres and has changed over time.[20, 21] Therefore not all iPCD patients were diagnosed according to current recommendations. We accounted for this by distinguishing three levels of diagnostic certainty: patients with definite PCD according to the ERS guidelines[19] with a hallmark ultrastructural defect identified by transmission electron microscopy (TEM) or pathogenic biallelic PCD genetic mutations; patients with probable PCD who had abnormal high-speed video microscopy findings or low nasal nitric oxide; and patients diagnosed on clinical grounds with an incomplete diagnostic algorithm. All patients had a clinical phenotype consistent with PCD, and other diagnoses had been excluded.

### Lung function

We calculated FEV_1_ and FVC z-scores adjusted for age, sex, height, and ethnicity using the GLI 2012 reference values.[22] We only included patients with at least three lung function measurements. Baseline FEV_1_ and FVC values were defined as the individuals’ first available measurement. Measurements done before the age of 6 years were excluded to improve comparability. We included only measurements from scheduled follow-up visits when patients were in a stable condition. We first computed average yearly individual trajectories of FEV_1_ and FVC by fitting a linear regression for each patient separately including the repeated lung function measurements and age. Then we classified individual FEV_1_ trajectories based on the natural variability of lung function in healthy children into three groups: those that improved over time (> 0.05 z-scores/year), remained stable (≤ 0.05 & ≥ -0.05 z-scores/year), and those that decreased (< - 0.05 z-scores/year).[23]

### Potential determinants of lung function and of lung function trajectories

We investigated associations of potential determinants on lung function by differentiating between effects on baseline lung function (the intercept) and effects on change of lung function over time (the slope). We included factors that had been described previously to affect lung function and were available from all centres: country of residence, sex, age at diagnosis of PCD, year of diagnosis, level of diagnostic certainty, organ laterality, age, body mass index (BMI) z-scores based on WHO reference values[24] at first lung function testing, and ultrastructural defect. All determinants except age were time-independent.

### Statistical analysis

In case of missing data we contacted PIs. If the missing data could not be retrieved, the record was excluded from analysis. For laterality defects, we coded patients with missing information as “situs not reported”. Because ultrastructural defects had not been assessed in all patients, we investigated these in a subgroup analysis.

We compared z-scores of FEV_1_ and FVC with the GLI reference values,[22] and calculated individual and average lung function trajectories over time using linear regression. Then we compared characteristics between patients with different slopes: improving, stable, and decreasing lung function. Last, we investigated associations of lung function with potential determinants in multivariable linear mixed effects regression models with a random intercept at the patient level. We tested for linearity of mean trajectory by including age as a linear and quadratic term and computed the p-value for the coefficient of the quadratic term. We also included a random slope to allow for individual differences in lung function change by age. To assess whether changes in FEV_1_ or FVC z-scores over time were modified by certain patient characteristics, we included interaction terms between age at lung function test and the potential determinants considered. We also tested for a possible interaction between age at diagnosis and country. We used likelihood ratio (LR) tests to calculate p-values for single variables and for interaction terms. For categorical variables, the largest group was chosen as reference category. We tested robustness of the parameter estimates across countries by running the regression model for single countries with more than 30 patients.

We investigated the effects of ultrastructural defects on lung function growth in a subgroup of children with available TEM results. Adjusting for the same determinants as before, we further included an interaction term between type of ultrastructural defect and age to assess whether lung function changes over time differed between ultrastructural defects. Ultrastructural defects were grouped as in recent publications[13, 25] into: (1) Normal TEM results; (2) Outer dynein arm defect (ODA); (3) Outer and inner dynein arm defects (IDA); (4) Microtubular disorganization defects, consisting of nexin link defects combined with IDA or tubular disorganization defects combined with IDA; (5) Central complex defects, defined as central pair defects or tubular transposition defects; (6)

Any other defect (i.e. isolated IDA, acilia or nexin link defects and tubular disorganization without IDA). Group 6 (any other defects) was excluded from the analysis. To assess the amount of variance explained by all the predictors in our model we calculated in this subgroup the marginal R^2^, i.e. the proportion of variance explained by the fixed factors, and conditional R^2^, .i.e. the proportion of variance explained by both the fixed and random factors, using the method by Nakagawa et al.[26] We then compared R^2^ of the models with and without addition of ultrastructural defects. All analyses were performed using R 3.5.1 (www.r-project.org), linear mixed models with the R-package lme4.

## Results

### Population characteristics

By March 2019, 20 centres had delivered cleaned and standardised longitudinal data for 4470 lung function tests from 486 individuals (Figure S1). In 130 patients, the first lung function test stemmed from the year PCD was diagnosed, in the others it was later, often because PCD had been diagnosed at pre-school age. The largest datasets came from Germany (60 patients), Denmark (59), Turkey (49) and Israel (41) (Table 1). Half of the patients were female (49%), mean age at diagnosis was 8.6 years, 17% had been diagnosed before 1999, 38% between 1999 and 2008 and 45% after 2009. The PCD diagnosis was classified as definite in 65%, probable in 28% and clinical in 7% of patients. Half of the patients (52%) had situs solitus, 37% situs inversus, 3% heterotaxia, and 8% unknown situs. Information on ultrastructural defects was available for 366 (75%). Dynein arm defects were identified in 213 patients, a central complex defect in 40, and microtubular disorganization in 28. Mean age at the first lung function test was 11 years, mean BMI z-score -0.04 (SD =1.27).

**Table 1.**
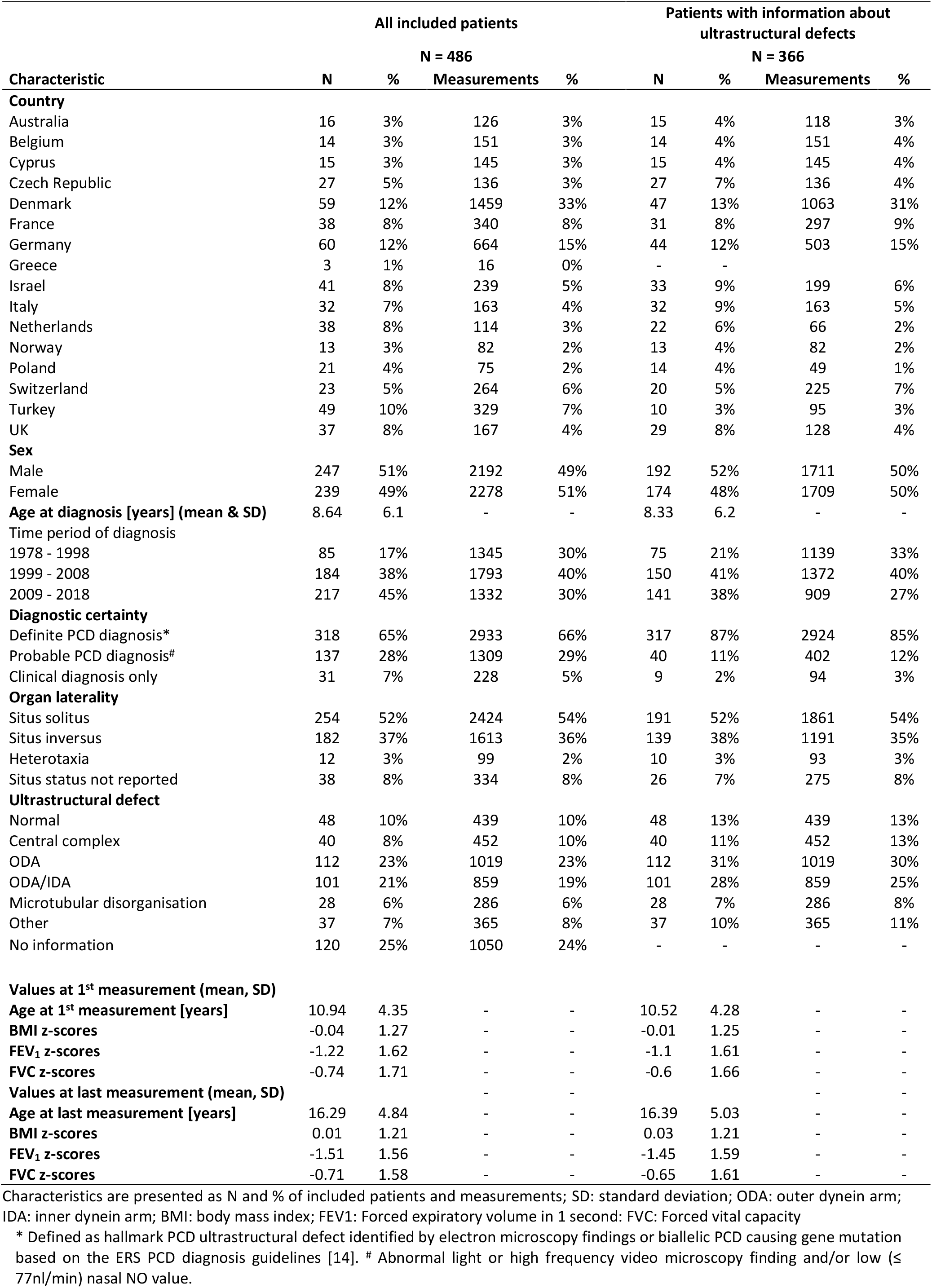
Demographic and baseline characteristics of patients with primary ciliary dyskinesia (PCD) (N=486)

### Lung function

Median number of lung function tests per participant was 6 (IQR: 4 - 11) and median follow-up time 4.14 years (IQR: 2.3 - 7.6). Lung function was below average already at baseline, with a mean (SD) FEV_1_ z-score of -1.22 (1.62) and mean (SD) FVC z-score of -0.74 (1.71) (Table 1, Figure 1 & S2). At the last test, mean (SD) FEV_1_ z-score was -1.51 (1.56) and mean (SD) FVC z-score -0.71 (1.58).

**Figure 1.**
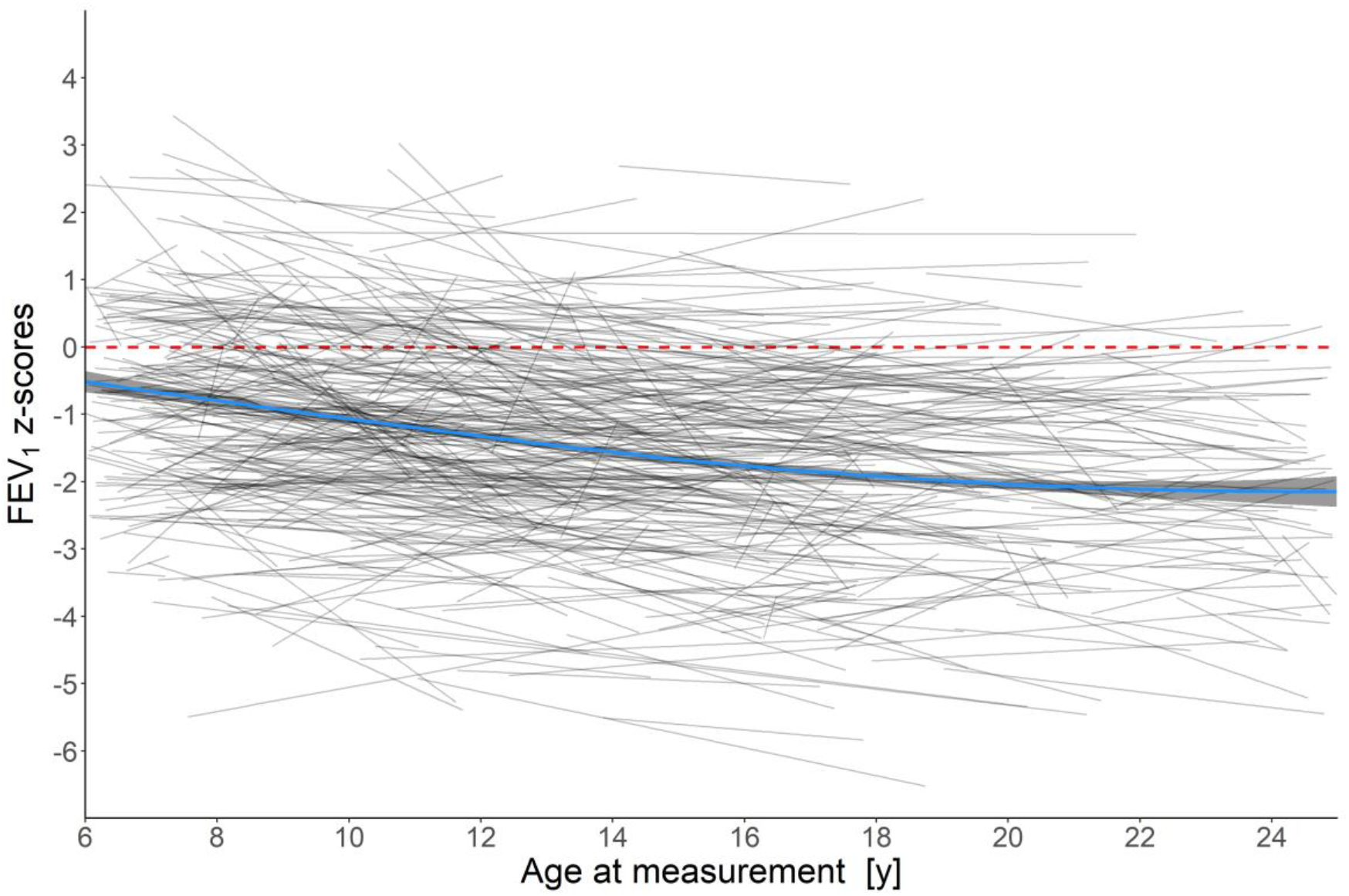
FEV_1_ trajectories during the lung growth period compared to Global Lung Function Initiative 2012 reference values. FEV_1_ (Forced expiratory volume in 1 second) is presented as z-score. A loess curve (blue line) was used to display the trajectory over time of all measurements and is plotted with a 95% confidence interval (shaded bands). Grey lines represent the individual linear trajectories of each patient included in the study. The dashed line shows the mean z-score of the normal population.

Average individual annual change in FEV1 was -0.06 z-scores (95% CI -0.072 to -0.057) and -0.03 z-scores (95% CI -0.040 to -0.023) for FVC. Lung function slopes differed between individuals: average yearly FEV1 improved by at least 0.05 z-scores in 21% of patients, remained stable in 40% and decreased in 39% (Table 2). Results from the regression analysis comparing FEV1 and FVC z-scores with GLI reference scores showed no evidence of a non-linear trend in lung function decline (table 3).

**Table 2.** Characteristics of patients with primary ciliary dyskinesia (PCD) from the international PCD cohort by FEV_1_ trajectories (lung function improving, stable, or decreasing over time) (N=486)

| FEV<sub>1</sub> trajectory* | Improving |  | Stable |  | Decreasing |  |
| --- | --- | --- | --- | --- | --- | --- |
|  | N | % | N | % | N | % |
| **Total** | 99 | 21% | 196 | 40% | 191 | 39% |
| **Country** |  |  |  |  |  |  |
| Australia | 3 | 19% | 5 | 31% | 8 | 50% |
| Belgium | 2 | 14% | 6 | 43% | 6 | 43% |
| Cyprus | 3 | 20% | 9 | 60% | 3 | 20% |
| Czech Republic | 6 | 22% | 13 | 48% | 8 | 30% |
| Denmark | 7 | 12% | 26 | 44% | 26 | 44% |
| France | 5 | 13% | 9 | 24% | 24 | 63% |
| Germany | 14 | 23% | 21 | 35% | 25 | 42% |
| Greece | 2 | 66% | 0 | 0% | 1 | 33% |
| Israel | 6 | 15% | 18 | 44% | 17 | 41% |
| Italy | 6 | 19% | 14 | 44% | 12 | 37% |
| Netherlands | 5 | 13% | 16 | 42% | 17 | 45% |
| Norway | 2 | 15% | 5 | 38% | 6 | 46% |
| Poland | 6 | 29% | 9 | 42% | 6 | 29% |
| Switzerland | 3 | 13% | 11 | 48% | 9 | 39% |
| Turkey | 21 | 43% | 16 | 33% | 12 | 24% |
| UK | 10 | 27% | 16 | 43% | 11 | 30% |
| **Sex** |  |  |  |  |  |  |
| Female | 49 | 21% | 98 | 41% | 92 | 38% |
| Male | 50 | 20% | 98 | 40% | 99 | 40% |
| **Age at diagnosis [years] (mean & SD)** | 9.89 | 5.74 | 8.59 | 6.46 | 8.06 | 5.84 |
| **Time period of diagnosis** |  |  |  |  |  |  |
| 1978 - 1998 | 11 | 13% | 33 | 39% | 41 | 48% |
| 1999 - 2008 | 35 | 19% | 73 | 40% | 76 | 41% |
| 2009 - 2018 | 53 | 24% | 90 | 41% | 74 | 34% |
| **Diagnostic certainty** |  |  |  |  |  |  |
| Definite PCD diagnosis | 55 | 17% | 137 | 43% | 126 | 40% |
| Probable PCD diagnosis | 38 | 28% | 49 | 36% | 50 | 36% |
| Clinical diagnosis only | 6 | 19% | 10 | 32% | 15 | 48% |
| **Organ laterality** |  |  |  |  |  |  |
| Situs solitus | 65 | 26% | 99 | 39% | 90 | 35% |
| Situs inversus | 26 | 14% | 79 | 43% | 77 | 42% |
| Heterotaxia | 2 | 16% | 5 | 42% | 5 | 42% |
| Situs status not reported | 6 | 16% | 13 | 34% | 19 | 50% |
| **Ultrastructural defect** |  |  |  |  |  |  |
| Normal | 10 | 20% | 19 | 40% | 19 | 40% |
| Central complex | 10 | 25% | 17 | 43% | 13 | 32% |
| ODA | 21 | 19% | 50 | 45% | 41 | 37% |
| ODA/IDA | 16 | 16% | 38 | 38% | 47 | 46% |
| Microtubular disorganisation | 3 | 11% | 14 | 50% | 11 | 39% |
| Other | 5 | 14% | 19 | 51% | 13 | 35% |
| No information | 34 | 28% | 39 | 33% | 47 | 39% |
| **BMI z-scores at 1st measurement (mean & SD)** | -0.2 | 1.25 | 0 | 1.32 | 0 | 1.23 |
| **BMI z-scores at last measurement (mean & SD)** | 0.02 | 1.2 | 0.13 | 1.18 | -0.13 | 1.24 |
| **FEV<sub>1</sub> z-scores at 1st measurement (mean & SD)** | -1.96 | 1.47 | -1.37 | 1.59 | -0.67 | 1.52 |
| **FEV<sub>1</sub> z-scores at last measurement (mean & SD)** | -0.98 | 1.38 | -1.34 | 1.57 | -1.96 | 1.52 |
| **FVC z-scores at 1st measurement (mean & SD)** | -1.43 | 1.72 | -0.94 | 1.69 | -0.17 | 1.53 |
| **FVC z-scores at last measurement (mean & SD)** | -0.45 | 1.49 | -0.58 | 1.59 | -0.97 | 1.59 |

SD: standard deviation; ODA: outer dynein arm; IDA: inner dynein arm; BMI: body mass index; FEV1: Forced expiratory volume in 1 second; FVC: Forced vital capacity
Yearly lung function change, improving: $> 0.05$ z-scores; stable: $\leq 0.05$ & $\geq -0.05$ z-scores; decreasing: $< -0.05$ z-scores. Data is presented with row percentages.

**Table 3.** FEV_1_ of patients with primary ciliary dyskinesia (PCD) from the international PCD cohort compared to Global Lung Function Initiative 2012 reference values (linear mixed effects regression, adjusting for all covariates).

| Variable | Estimate | FEV<sub>1</sub> |  | p-value<sup>¶</sup> |
| --- | --- | --- | --- | --- |
|  |  | 95% CI |  |  |
| **Intercept (Lung function at age 6 years for “reference patient”) \*** | -1.26 | -2.19 | -0.3 |  |
| **Age at measurement** | -0.04 | -0.12 | 0.04 |  |
| **Country (Ref: Germany)** |  |  |  | <0.01 |
| Australia | -0.34 | -1.26 | 0.59 |  |
| Belgium | -0.20 | -1.03 | 0.64 |  |
| Cyprus | -1.19 | -2.14 | -0.23 |  |
| Czech Republic | -0.95 | -1.68 | -0.22 |  |
| Denmark | 0.21 | -0.42 | 0.84 |  |
| France | 0.14 | -0.65 | 0.93 |  |
| Greece | 0.92 | -2.35 | 4.19 |  |
| Israel | 0.11 | -0.71 | 0.93 |  |
| Italy | -0.28 | -1.02 | 0.47 |  |
| Netherlands | 1.63 | 0.84 | 2.43 |  |
| Norway | -0.31 | -1.24 | 0.63 |  |
| Poland | -0.86 | -1.70 | -0.03 |  |
| Switzerland | 0.13 | -0.63 | 0.89 |  |
| Turkey | -1.76 | -2.46 | -1.06 |  |
| UK | -1.09 | -1.90 | -0.28 |  |
| **Sex (Ref: male)** |  |  |  | 0.37 |
| Female | -0.03 | -0.33 | 0.27 |  |
| **Age at diagnosis** | -0.01 | -0.05 | 0.02 | 0.21 |
| **Diagnostic period** | 0.02 | -0.005 | 0.05 | 0.26 |
| **Diagnostic certainty (Ref: definite PCD diagnosis)** |  |  |  | 0.49 |
| Probable PCD diagnosis | -0.04 | -0.43 | 0.35 |  |
| Clinical diagnosis only | 0.38 | -0.27 | 1.03 |  |
| **Laterality defects (Ref: situs solitus)** |  |  |  | 0.59 |
| Situs Ambiguous | 0.42 | -0.53 | 1.36 |  |
| Situs Inversus | 0.33 | -0.001 | 0.66 |  |
| Situs Unknown | 0.15 | -0.54 | 0.85 |  |
| **BMI** | 0.32 | 0.28 | 0.37 | <0.01 |
| **Change of lung function over time<sup>#</sup>** |  |  |  |  |
| **Country (Ref: Germany)** |  |  |  | <0.01 |
| Australia | -0.02 | -0.10 | 0.07 |  |
| Belgium | 0.02 | -0.05 | 0.10 |  |
| Cyprus | 0.07 | -0.01 | 0.15 |  |
| Czech Republic | 0.04 | -0.03 | 0.11 |  |
| Denmark | -0.01 | -0.07 | 0.05 |  |
| France | -0.05 | -0.12 | 0.03 |  |
| Greece | 0.01 | -0.29 | 0.32 |  |
| Israel | -0.05 | -0.13 | 0.03 |  |
| Italy | -0.03 | -0.10 | 0.04 |  |
| Netherlands | -0.14 | -0.24 | -0.04 |  |
| Norway | -0.02 | -0.12 | 0.09 |  |
| Poland | 0.04 | -0.05 | 0.13 |  |
| Switzerland | -0.03 | -0.10 | 0.05 |  |
| Turkey | 0.11 | 0.04 | 0.18 |  |
| UK | 0.03 | -0.04 | 0.11 |  |
| **Sex (Ref: male)** |  |  |  | 0.36 |
| Female | -0.01 | -0.04 | 0.02 |  |
| **Age at diagnosis** | 0.003 | 0.00 | 0.01 | 0.08 |
| **Diagnostic period** | -0.001 | -0.004 | 0.001 | 0.32 |
| **Diagnostic certainty (Ref: definite PCD diagnosis)** |  |  |  | 0.92 |
| Probable PCD diagnosis | -0.01 | -0.04 | 0.03 |  |
| Clinical diagnosis only | -0.001 | -0.07 | 0.07 |  |
| **Laterality defects (Ref: situs solitus)** |  |  |  | 0.52 |
| Situs Ambiguous | -0.01 | -0.11 | 0.08 |  |
| Situs Inversus | -0.02 | -0.05 | 0.01 |  |
| Situs Unknown | -0.02 | -0.08 | 0.04 |  |
| **BMI** | 0.02 | 0.01 | 0.02 | <0.01 |

FEV<sub>1</sub>: Forced expiratory volume in 1 second; BMI: body mass index
¶ Likelihood ratio test p-value indicating whether the characteristic explains differences in FEV<sub>1</sub> within the study population.
\*The Intercept describes the FEV<sub>1</sub> of a reference patient at 6 years, who is male, from Germany, with a BMI z-score of 0, diagnosed at birth (age = 0) in 1978, with a definite PCD diagnosis. Categorical variables describe the change from the reference category, while continuous variables describe the change from the reference patient for each unit of increase.
\* Defined as hallmark PCD ultrastructural defect identified by electron microscopy findings or biallelic PCD causing gene mutation based on the ERS PCD diagnosis guidelines [14]. # Abnormal light or high frequency video microscopy finding and/or low ( $\leq 77$ nl/min) nasal NO value.

### Determinants of lung function

Lung function differed by countries (Table 3 & S1, Figure 2 & S3). FEV_1_ was reduced at the age of 6 years in most countries compared to GLI reference values, except the Netherlands. FEV_1_ z-scores were lowest in Turkey (−3.02), Cyprus (−2.45) and the UK (−2.35), and highest in the Netherlands (0.37) and Denmark (−1.05). The change in FEV_1_ over time was also variable; lung function deteriorated over time in most countries. Yearly decline of FEV_1_ z-scores was highest in the Netherlands (−0.14), Israel (−0.05) and France (−0.05). FEV_1_ z-scores increased over time in Turkey (0.11), Cyprus (0.07) and Poland (0.04). For FVC (Table S1 and Figure S3), the results were similar to FEV_1_. FVC also decreased over time in most countries, except for Turkey (0.08), with the steepest decline in the Netherlands (−0.26).

**Figure 2.**
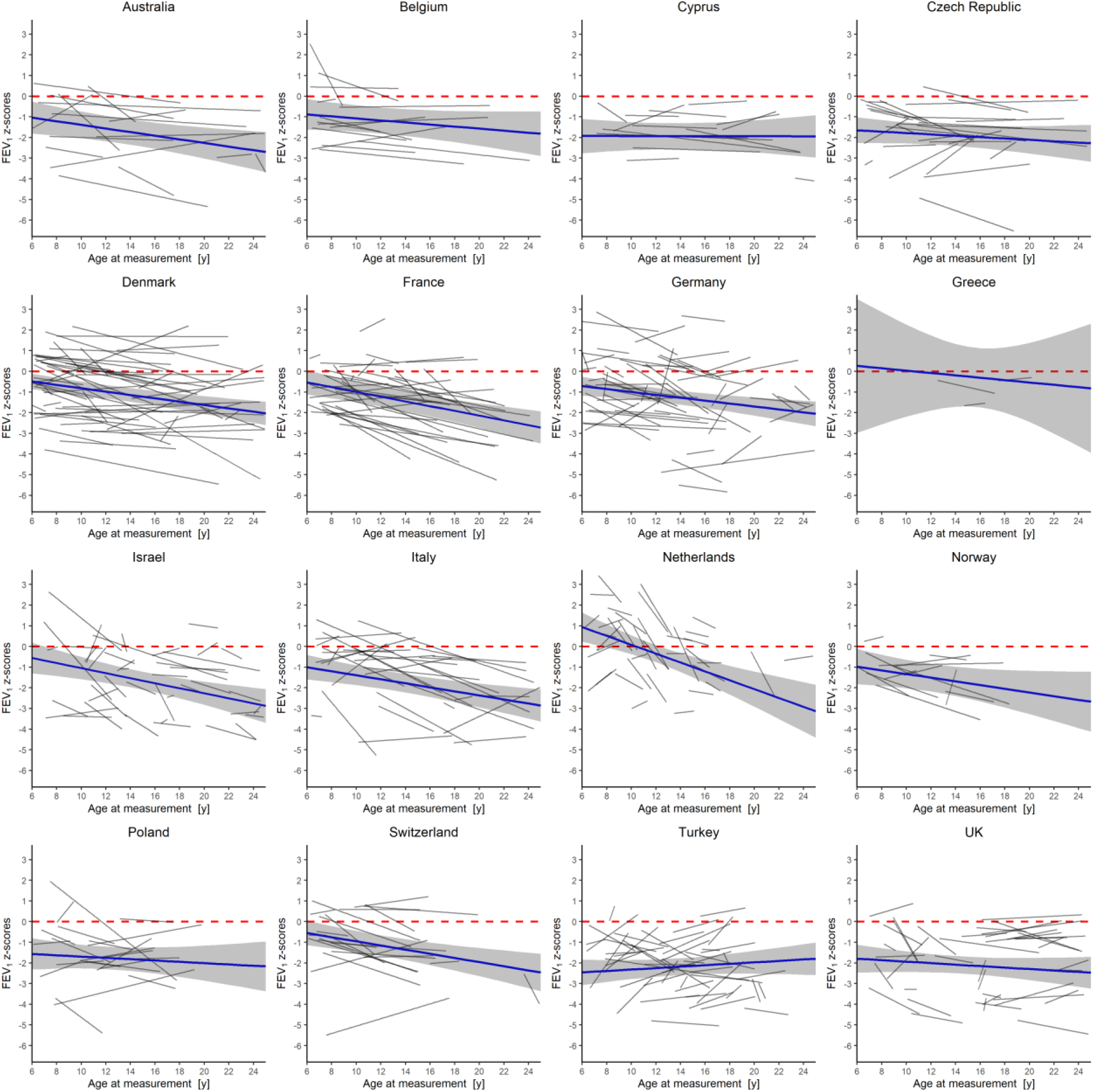
FEV_1_ trajectories of PCD patients in different countries compared to Global Lung Function Initiative (GLI) 2012 reference values. Estimated linear individual trajectories are shown as black lines (based on individual linear regressions), estimate linear mean trajectories (based on mixed linear regression model for subgroup) as blue lines, and point-wise 95% confidence intervals as grey shaded areas. The dashed line shows the mean z-score of the normal population (GLI 2012). FEV1: Forced expiratory volume in 1 second

BMI was positively associated with FEV_1_ and FVC. Patients with higher BMI had a better lung function at baseline and improved more over time (Table 3 & S1, Figure 3). Baseline FEV_1_ was 0.32 z-scores higher for each unit of higher BMI z-score. Yearly change of FEV_1_ increased by 0.02 z-scores for each unit of z-score increase in BMI. Similarly, baseline FVC and increase of FVC over time were higher in individuals with higher BMI. BMI remained a determinant for lung function when running separate regression model for countries with at least 30 patients (table S3).

Ultrastructural defects were also associated with lung function (Table 4). Patients with microtubular disorganization defects had the worst baseline FEV_1_ z-scores (−1.75), followed by patients with central complex defects (−1.38) and ODA defects (−1.27). Patients with non-diagnostic TEM (−0.91) or combined ODA & IDA defects (−0.60) had the best lung function at baseline. Yearly changes of FEV_1_ also varied between groups, with the steepest decline in patients with ODA & IDA defects (−0.05), and the most favourable course in patients with central complex defects. FVC varied similarly with ultrastructural defects (Table S2), but the evidence was less strong as for FEV_1_. The amount of variability explained by the model changed only marginally when we additionally included ultrastructural defects into the model: the marginal R^2^ changed from 0.211 to 0.237 and the conditional R^2^ from 0.817 to 0.819.

**Table 4.**
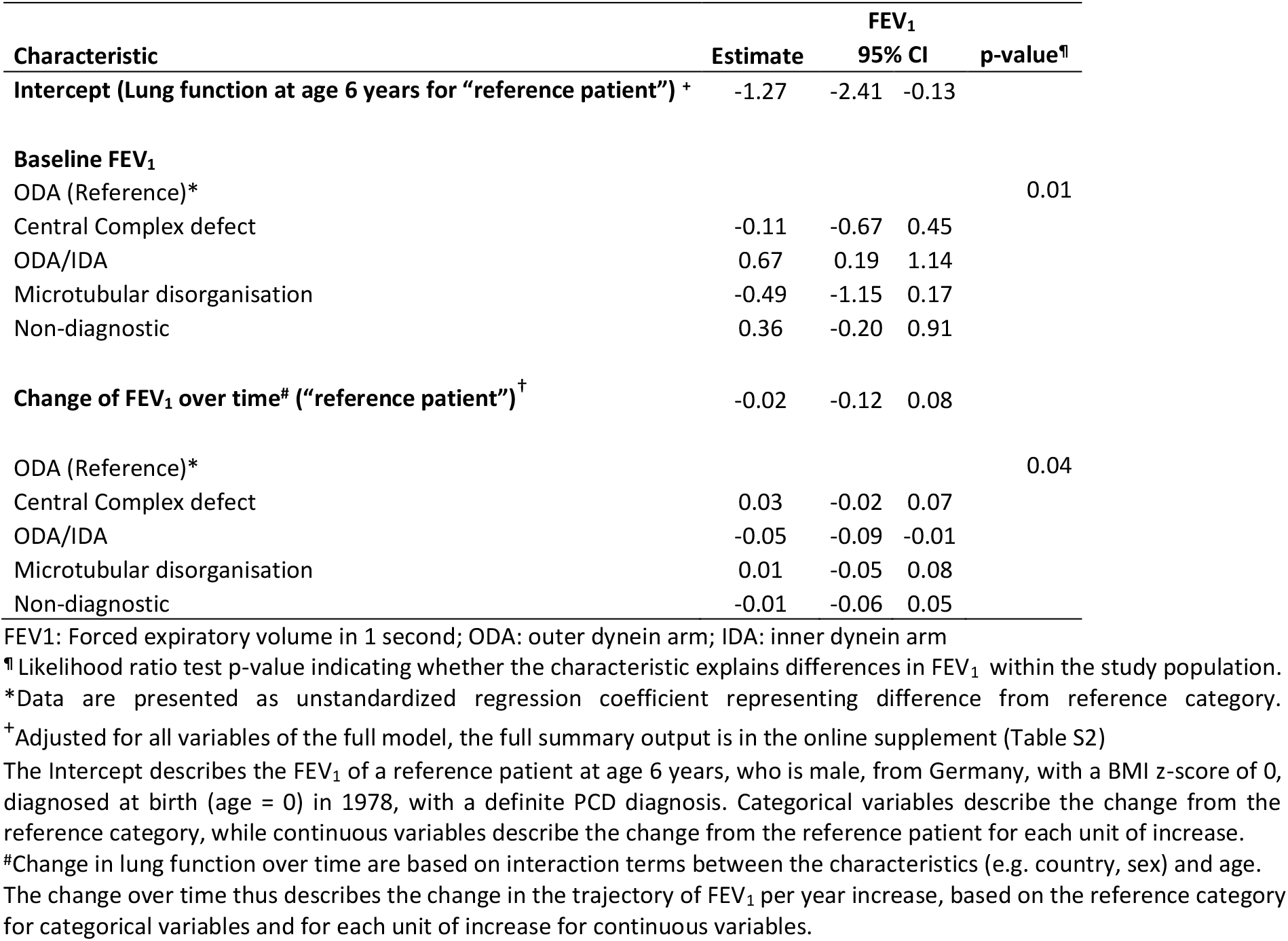
FEV_1_ in patients with primary ciliary dyskinesia (PCD), by ultrastructural defect (N = 366).

| Characteristic | Estimate | FEV<sub>1</sub> |  | p-value<sup>¶</sup> |
| --- | --- | --- | --- | --- |
|  |  | 95% CI |  |  |
| **Intercept (Lung function at age 6 years for “reference patient”) <sup>+</sup>** | -1.27 | -2.41 | -0.13 |  |
| **Baseline FEV<sub>1</sub>** |  |  |  |  |
| ODA (Reference)* |  |  |  | 0.01 |
| Central Complex defect | -0.11 | -0.67 | 0.45 |  |
| ODA/IDA | 0.67 | 0.19 | 1.14 |  |
| Microtubular disorganisation | -0.49 | -1.15 | 0.17 |  |
| Non-diagnostic | 0.36 | -0.20 | 0.91 |  |
| **Change of FEV<sub>1</sub> over time<sup>#</sup> (“reference patient”) <sup>†</sup>** | -0.02 | -0.12 | 0.08 |  |
| ODA (Reference)* |  |  |  | 0.04 |
| Central Complex defect | 0.03 | -0.02 | 0.07 |  |
| ODA/IDA | -0.05 | -0.09 | -0.01 |  |
| Microtubular disorganisation | 0.01 | -0.05 | 0.08 |  |
| Non-diagnostic | -0.01 | -0.06 | 0.05 |  |

FEV<sub>1</sub>: Forced expiratory volume in 1 second; ODA: outer dynein arm; IDA: inner dynein arm
<sup>¶</sup> Likelihood ratio test p-value indicating whether the characteristic explains differences in FEV<sub>1</sub> within the study population.
\*Data are presented as unstandardized regression coefficient representing difference from reference category.
<sup>†</sup> Adjusted for all variables of the full model, the full summary output is in the online supplement (Table S2)
The Intercept describes the FEV<sub>1</sub> of a reference patient at age 6 years, who is male, from Germany, with a BMI z-score of 0, diagnosed at birth (age = 0) in 1978, with a definite PCD diagnosis. Categorical variables describe the change from the reference category, while continuous variables describe the change from the reference patient for each unit of increase.
<sup>#</sup> Change in lung function over time are based on interaction terms between the characteristics (e.g. country, sex) and age.
The change over time thus describes the change in the trajectory of FEV<sub>1</sub> per year increase, based on the reference category for categorical variables and for each unit of increase for continuous variables.

We did not find evidence that FEV_1_ and FVC z-scores differed by sex, age at diagnosis, year of diagnosis, laterality defects, or level of diagnostic certainty (Table 3 & S1).

## Discussion

This large international study of patients with PCD found that lung function, already reduced by the age of 6 years, declined further during childhood, adolescence, and early adulthood with an overall negative trend in z-scores. There was much heterogeneity between countries and individuals. Overall, FEV_1_ z-scores decreased over time in 39% of children, remained stable in 40% and improved in 21%. Type of ultrastructural defect was marginally associated and BMI more strongly associated with lung function at baseline and changes in lung function over time.

With 4470 lung function tests from 486 individuals this study is by far the largest showing lung function trajectories in children, adolescents, and young adults with PCD. Most other longitudinal studies combined children and older adults, not allowing to distinguish the growth period in childhood from functional decline in adulthood.[8, 25] The large study population made it possible to compare countries and ultrastructural phenotypes and test for association with possible risk factors. A limitation of the study was that we lacked information on other factors that can influence lung function associated with bronchiectasis from any cause such as frequency of exacerbations, colonizing organisms, and patterns of care. Prospective cohort studies using standardised protocols would help to clarify the relative contribution of these factors to the rate of lung function change for patients with PCD.[27]

The wide variation in lung function between countries confirms previous cross-sectional data which included adults.[15] We observed that countries with poor lung function at baseline, such as Poland and Turkey, tended to have a less steep decline than countries with more favourable baseline values such as the Netherlands, Denmark, or France. Reasons for this are unclear but could include regression to the mean and differences in management of PCD. It is possible that patients who presented with a poor lung function at diagnosis were offered a strict physiotherapy and antibiotics management and were monitored more closely than patients who were better off at diagnosis. Also, length of follow-up time differed between countries, which could have affected the estimation of the lung function trajectories. In the Netherlands and France where the decline in lung function was steep, the mean lung function trajectory was estimated based on short individual follow-ups, while in countries with longer follow-ups, the decline in lung function was less steep such as Denmark or Belgium. We modelled mean trajectories as linear although they may be non-linear. However, in formal testing including a quadratic term, we found no evidence for non-linearity. It is also possible that the GLI reference values might not be applicable to all countries e.g. the Netherlands. However, if that was true, we would expect regional differences in baseline lung function, but the effect on slopes should be minor. Despite the differences between countries, results from the individual country models (Table S3) highlight the robustness of our findings.

Can early diagnosis prevent disease progression? This has been suggested by previous studies,[14, 28-30] while the Danish study and ours found no association.[8] As we still lack robust evidence for the best treatments for PCD, management is based on experience from other lung diseases such as CF. Therefore, PCD diagnosis might not have led to a change in management, as patients may have been treated for chronic lung disease already before PCD was diagnosed. PCD-specific randomised controlled trials will provide more specific treatment recommendations.[31, 32]

When considering the different phases of lung function over a lifetime,[9, 10] what have we learntã Our study and the North American one suggest that lung function growth during childhood is impaired.[13] The fact that lung function was already low at age six could be the result of poorer lung growth at preschool age or a reduced lung function at birth, or it could be a consequence of neonatal respiratory problems such as atelectasis or neonatal pneumonia. This can only be investigated using infant lung function testing in children with PCD diagnosed as neonates. Studies based on mainly adult patients reported larger declines (−0.89%/year in Italy)[8, 25] suggesting that also the decline of lung function in adulthood is accelerated. The fact that FEV_1_ z-scores decreased over time in all studies reflects long-term irreversible lung damage such as bronchiectasis and lung remodelling after recurrent severe infections.[33] On the positive side, the heterogeneity between patients that we followed suggests that many changes are reversible after initiation of regular physiotherapy and antibiotics, such as mucus plugging, bronchial wall thickening and temporary atelectasis.

Genetic and ultrastructural differences have been offered as an explanation for variable lung function trajectories in smaller studies.[8, 13, 34] Particularly poor lung function was seen in patients with microtubular defects.[6, 13, 29, 35] A UK study in 82 adults reported an FEV_1_ decline of 0.75 percent predicted in patients with microtubular defects compared to −0.51 percent predicted in patients with ODA or combined ODA and IDA defects, and −0.13 percent predicted in patients with normal or inconclusive TEM. In the North American study, annual change in FEV_1_ percent predicted deviated from normal only in patients with central apparatus and microtubular defects (−1.11 percent predicted) and not in patients with dynein arm defects or normal TEM [13]. Also in Italy, the steepest FEV_1_ decline was in patients with central apparatus and microtubular defects.[25] We did not have genetic data available for all patients to examine specific mutations. We found microtubular defects to be associated with the worst baseline lung function, but further course did not differ. The total variability explained by the regression model (marginal R^2^) increased only slightly when we included the ultrastructural defects, suggesting that ultrastructural differences explain only a small part of the heterogeneity.

BMI was associated with baseline lung function and with further change. This confirms previous cross-sectional data.[14, 15, 36] The effect of poor nutrition on lung function is known for patients with other respiratory diseases[37-39] and dietary support should be provided when required. In contrast to our previous cross-sectional analysis where we found that FEV1 and FVC z-scores were lower in females than males, we found no evidence of an association of lung function and sex.

However, here we included only children and young adults, while the previous study included older adults. In healthy individuals, female sex hormones might inhibit the function of the mucociliary apparatus.[40] As hormonal differences manifest after puberty, this could explain the discrepant results.

In conclusion, this large international study found considerable heterogeneity in lung function trajectories of children, adolescents, and young adults with PCD and a wide variation between countries. Lung function was low in 6-year-olds and declined further throughout the lung growth period despite treatment. It is essential to develop PCD specific treatment strategies to improve prognosis.

## Data Availability

The study database is not be shared publicly, given the confidentiality limitations to protect sensitive data.
Researchers wanting to use the data can propose a topic and a concept sheet describing the planned analyses and publication. All concept sheets have to be approved by all centres contributing data to the proposed analysis under question.

## Supplementary display items

**Figure S1.**
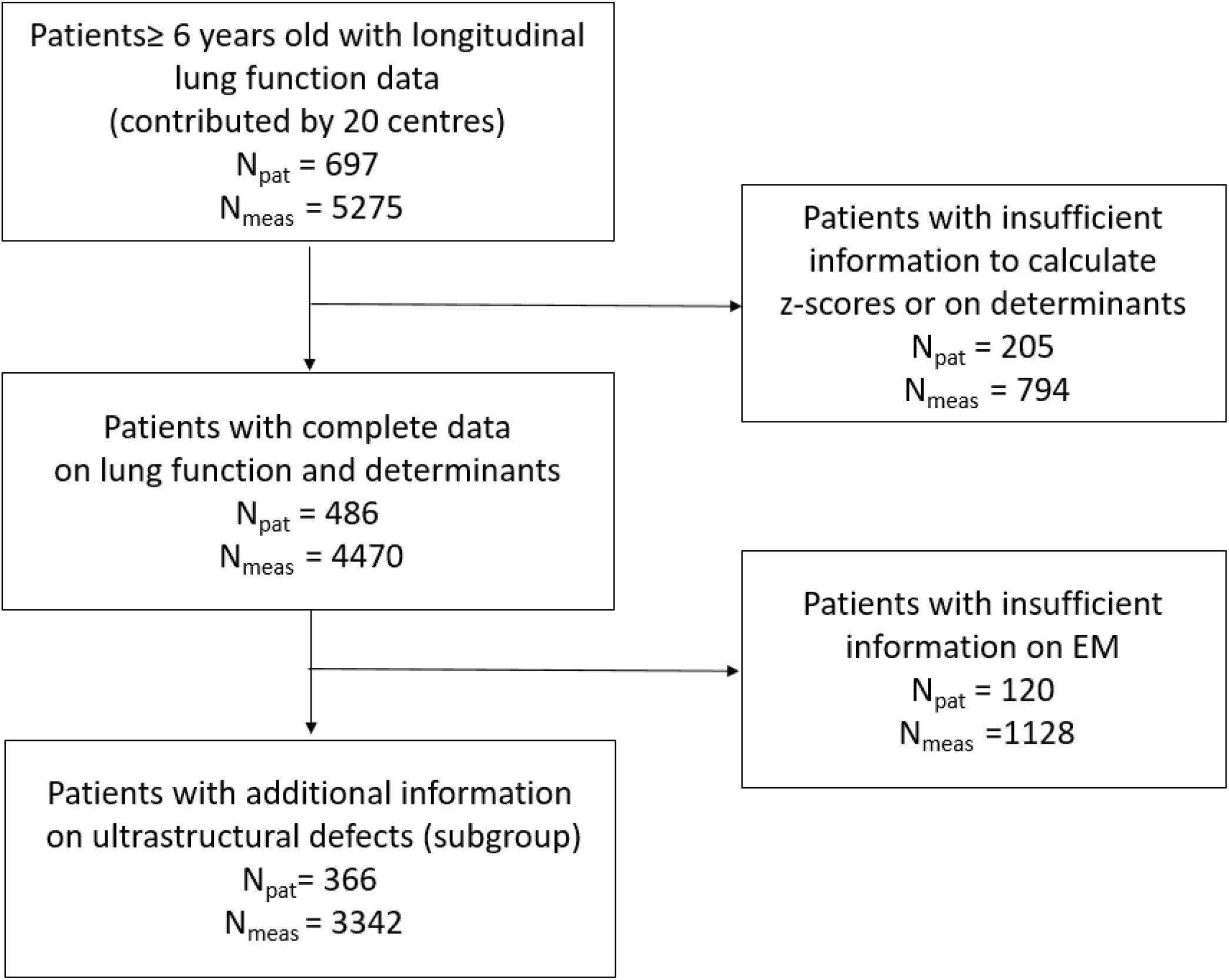
Flow chart showing the patients and respective lung function measurements included for the different analyses. Npat: number of patients; Nmeas: number of measurements; EM: electron microscopy

**Figure S2.**
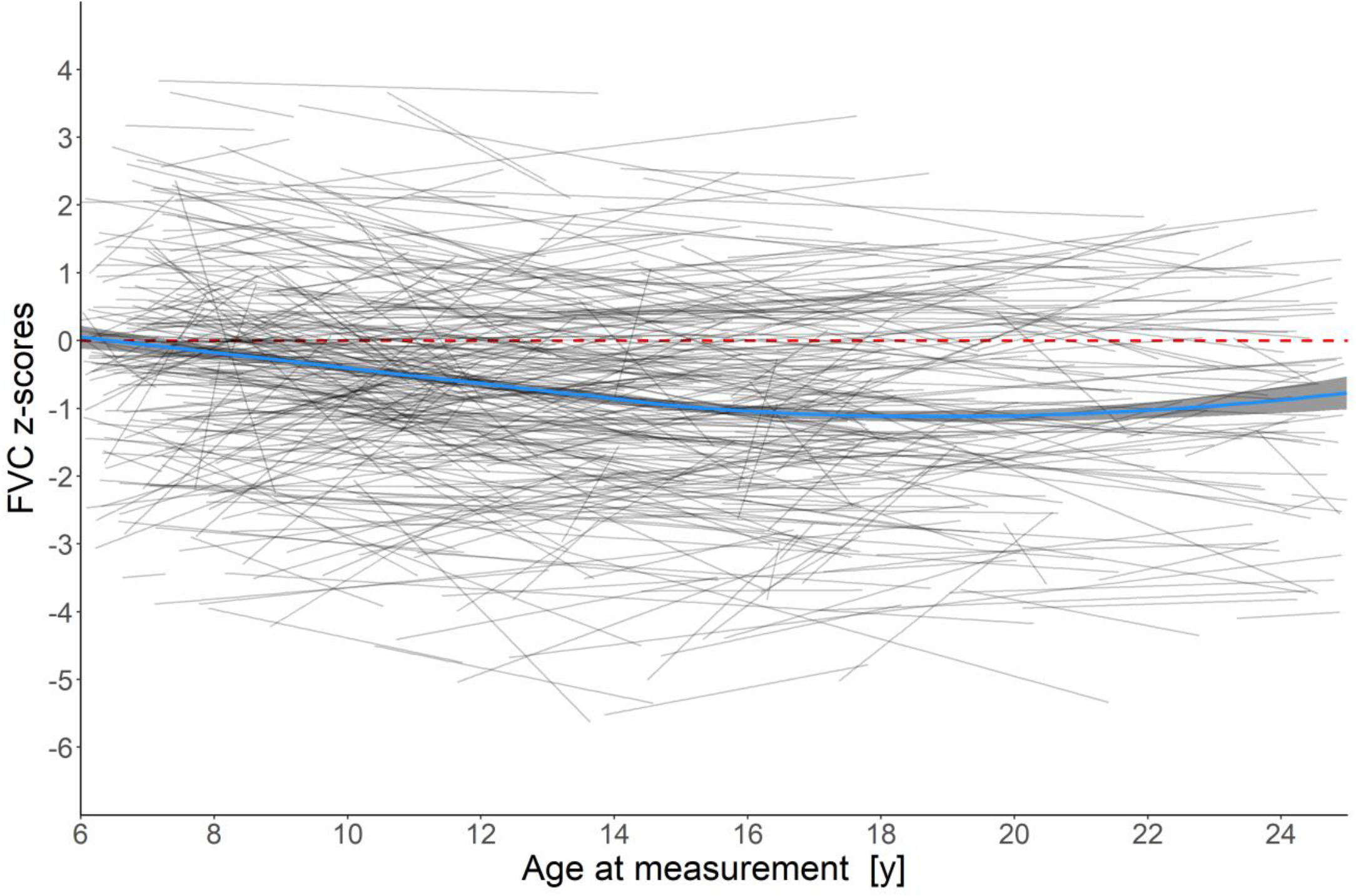
FVC trajectories during the lung growth period compared to Global Lung Function Initiative 2012 reference values. FVC (Forced vital capacity) is presented as z-score. A loess curve (blue line) was used to display the trajectory over time of all measurements and is plotted with a 95% confidence interval (shaded bands). Grey lines represent the individual linear trajectories of each patient included in the study. The dashed line shows the mean z-score of the normal population

**Table S1.** FVC of patients with primary ciliary dyskinesia (PCD) from the international PCD cohort, compared to Global Lung Function Initiative 2012 reference values (linear mixed effects regression, adjusting for all covariates).

| Variable | Estimate | FVC |  | p-value<sup>¶</sup> |
| --- | --- | --- | --- | --- |
|  |  | 95% CI |  |  |
| **Intercept** (Lung function at age 6 years for “reference patient”) + | -1.10 | -2.07 | -0.13 |  |
| **Age at measurement** | 0.02 | -0.07 | 0.11 |  |
| **Country** (Ref: Germany) |  |  |  | <0.01 |
| Australia | 0.06 | -0.90 | 1.02 |  |
| Belgium | 0.76 | -0.10 | 1.62 |  |
| Cyprus | -0.82 | -1.81 | 0.17 |  |
| Czech Republic | -0.73 | -1.49 | 0.03 |  |
| Denmark | 1.09 | 0.43 | 1.73 |  |
| France | 0.80 | -0.02 | 1.63 |  |
| Greece | 1.09 | -2.46 | 4.64 |  |
| Israel | 0.36 | -0.49 | 1.22 |  |
| Italy | 0.52 | -0.25 | 1.30 |  |
| Netherlands | 2.98 | 2.16 | 3.81 |  |
| Norway | 0.38 | -0.59 | 1.36 |  |
| Poland | -0.01 | -0.88 | 0.87 |  |
| Switzerland | 0.52 | -0.26 | 1.31 |  |
| Turkey | -1.78 | -2.51 | -1.05 |  |
| UK | -0.20 | -1.05 | 0.65 |  |
| **Sex** (Ref: male) |  |  |  | 0.79 |
| Female | -0.11 | -0.42 | 0.20 |  |
| **Age at diagnosis** | -0.02 | -0.05 | 0.01 | 0.47 |
| **Diagnostic period** | 0.01 | -0.02 | 0.04 | 0.49 |
| **Diagnostic certainty** (Ref: definite PCD diagnosis) |  |  |  | 0.30 |
| Probable PCD diagnosis | -0.12 | -0.52 | 0.29 |  |
| Clinical diagnosis only | 0.33 | -0.35 | 1.01 |  |
| **Laterality defects** (Ref: Situs solitus) |  |  |  | 0.40 |
| Situs Ambiguous | 0.16 | -0.83 | 1.15 |  |
| Situs Inversus | 0.30 | -0.05 | 0.64 |  |
| Situs Unknown | 0.00 | -0.74 | 0.72 |  |
| **BMI** | 0.40 | 0.35 | 0.45 | <0.01 |
| **Change of lung function over time** |  |  |  |  |
| **Country** (Ref: Germany) |  |  |  | <0.01 |
| Australia | -0.05 | -0.13 | 0.04 |  |
| Belgium | -0.10 | -0.18 | -0.02 |  |
| Cyprus | -0.04 | -0.12 | 0.05 |  |
| Czech Republic | -0.02 | -0.09 | 0.06 |  |
| Denmark | -0.05 | -0.12 | 0.01 |  |
| France | -0.10 | -0.18 | -0.03 |  |
| Greece | -0.08 | -0.41 | 0.25 |  |
| Israel | -0.09 | -0.17 | -0.01 |  |
| Italy | -0.09 | -0.17 | -0.02 |  |
| Netherlands | -0.26 | -0.37 | -0.16 |  |
| Norway | -0.07 | -0.18 | 0.04 |  |
| Poland | -0.04 | -0.14 | 0.06 |  |
| Switzerland | -0.06 | -0.14 | 0.02 |  |
| Turkey | 0.08 | 0.01 | 0.16 |  |
| UK | -0.05 | -0.13 | 0.04 |  |
| **Sex** (Ref: male) |  |  |  | 0.61 |
| Female | 0.01 | -0.02 | 0.04 |  |
| **Age at diagnosis** | 0.002 | -0.001 | 0.005 | 0.24 |
| **Diagnostic period** | -2.12 | -0.003 | 0.003 | 0.99 |
| **Diagnostic certainty** (Ref: definite PCD diagnosis) |  |  |  | 0.21 |
| Probable PCD diagnosis | -0.01 | -0.05 | 0.03 |  |
| Clinical diagnosis only | -0.07 | -0.14 | 0.01 |  |
| **Laterality defects** (Ref: situs solitus) |  |  |  | 0.73 |
| Situs Ambiguous | -0.004 | -0.10 | 0.09 |  |
| Situs Inversus | -0.01 | -0.04 | 0.02 |  |
| Situs Unknown | -0.04 | -0.10 | 0.03 |  |
| **BMI** | 0.01 | -0.005 | 0.02 | 0.01 |

FVC: Forced vital capacity; BMI: body mass index
¶ Likelihood ratio test p-value indicating whether the characteristic explains differences in FVC within the study population.
\*The Intercept describes the FVC of a reference patient at 6 years, who is male, from Germany, with a BMI z-score of 0, diagnosed at age 6 years in 1978, with a definite PCD diagnosis. Categorical variables describe the change from the reference category, while continuous variables describe the change from the reference patient for each unit of increase. \* Defined as hallmark PCD ultrastructural defect identified by electron microscopy findings or biallelic PCD causing gene mutation based on the ERS PCD diagnosis guidelines [14]. # Abnormal light or high frequency video microscopy finding and/or low ( $\leq 77$ nl/min) nasal NO value. Change of lung function over time describes the change in the trajectory of FEV<sub>1</sub> per year increase, based on the reference category for categorical variables and for each unit of increase for continuous variables.

**Figure S3.**
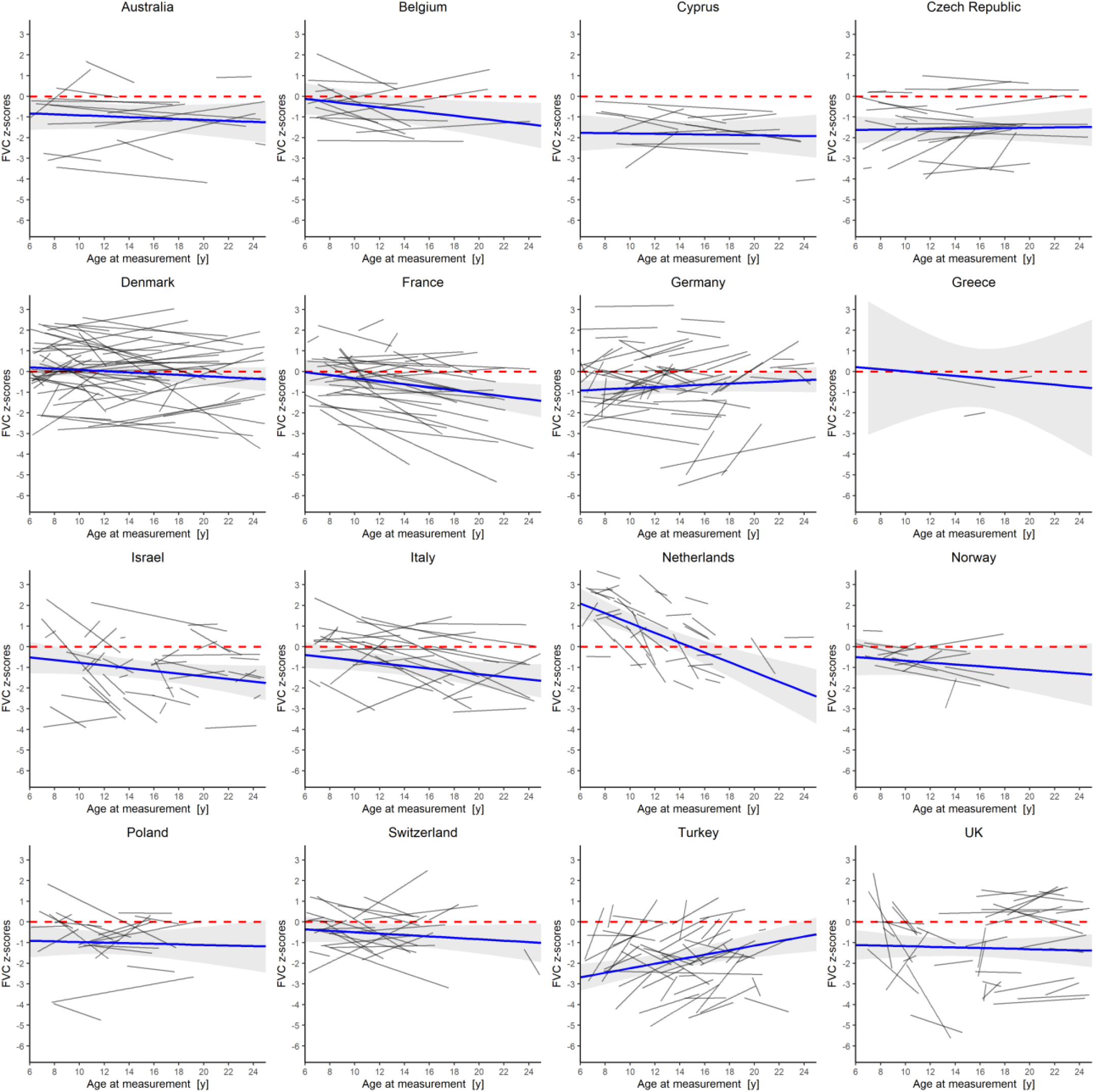
FVC trajectories of PCD patients in different countries compared to Global Lung Function Initiative (GLI) 2012 reference values. Individual trajectories are shown as black lines, marginal effects (estimated regression line for subgroup) as blue lines, 95% confidence intervals as grey shaded areas. The dashed line shows the mean z-score of the normal population (GLI 2012). FVC: Forced vital capacity

**Table S2.**
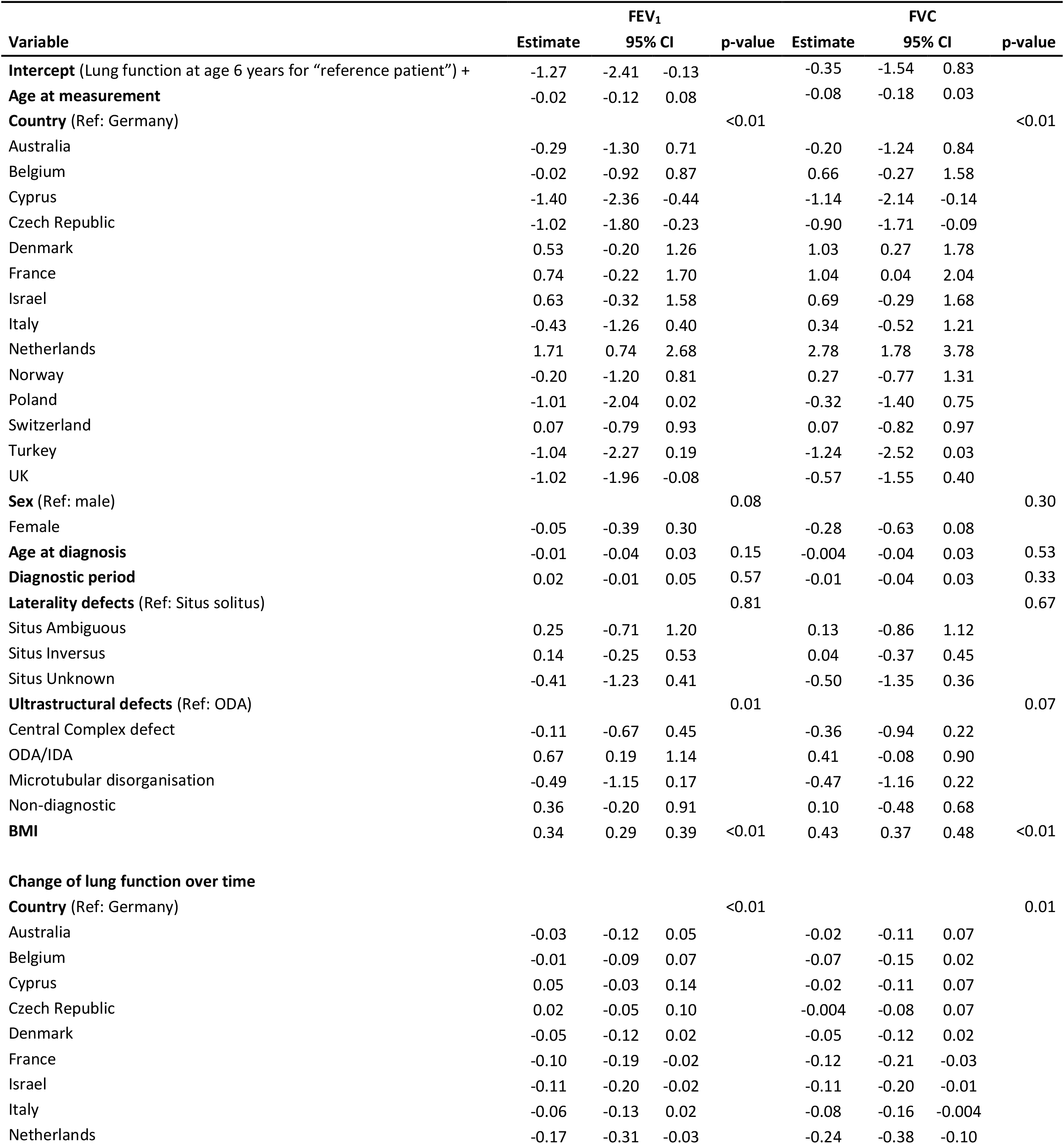
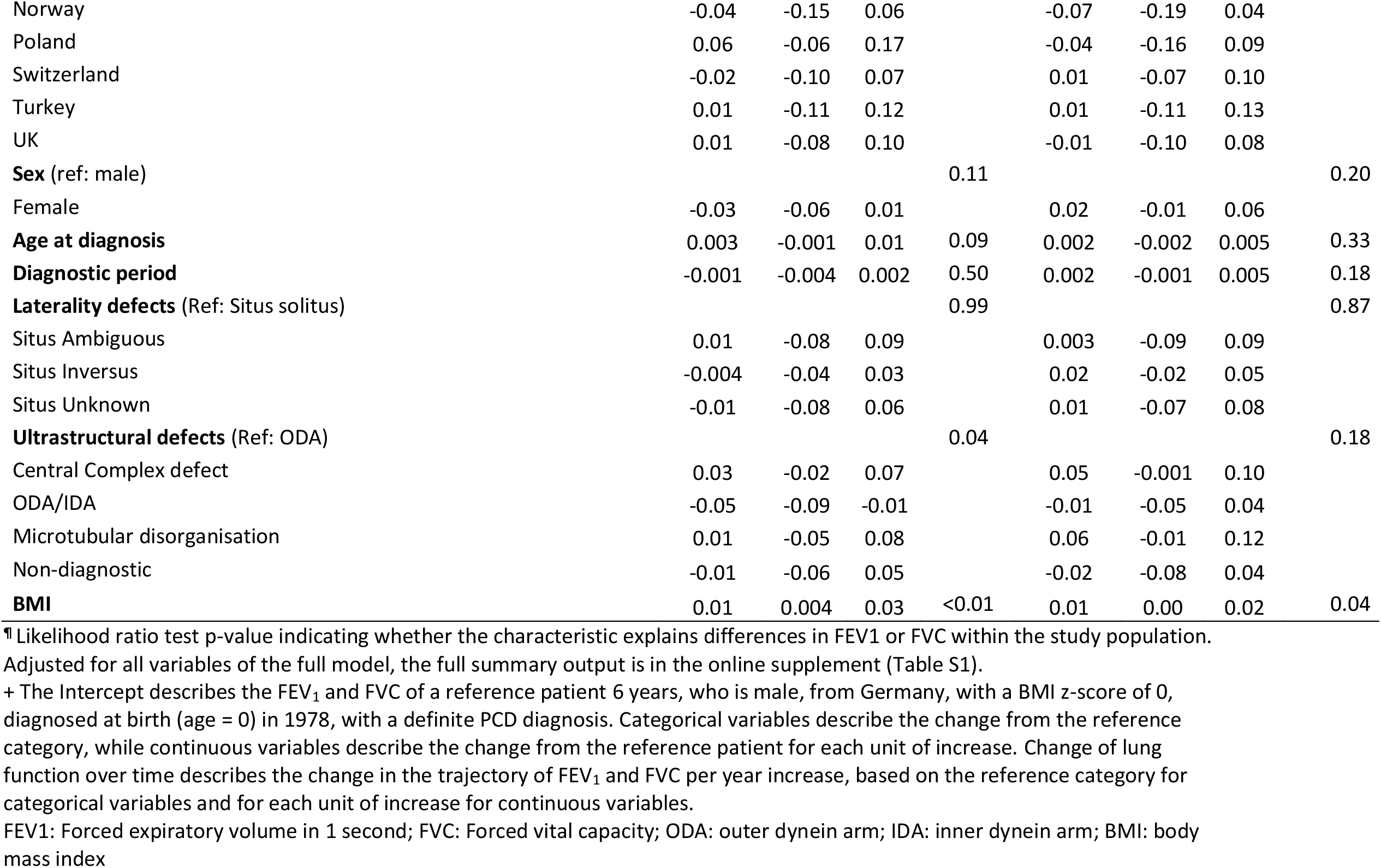
FEV1 and FVC of patients with primary ciliary dyskinesia (PCD) from the international PCD cohort with available ultrastructural defect information compare to Global Lung Function Initiative 2012 reference values (N = 366).

**Table S3.**
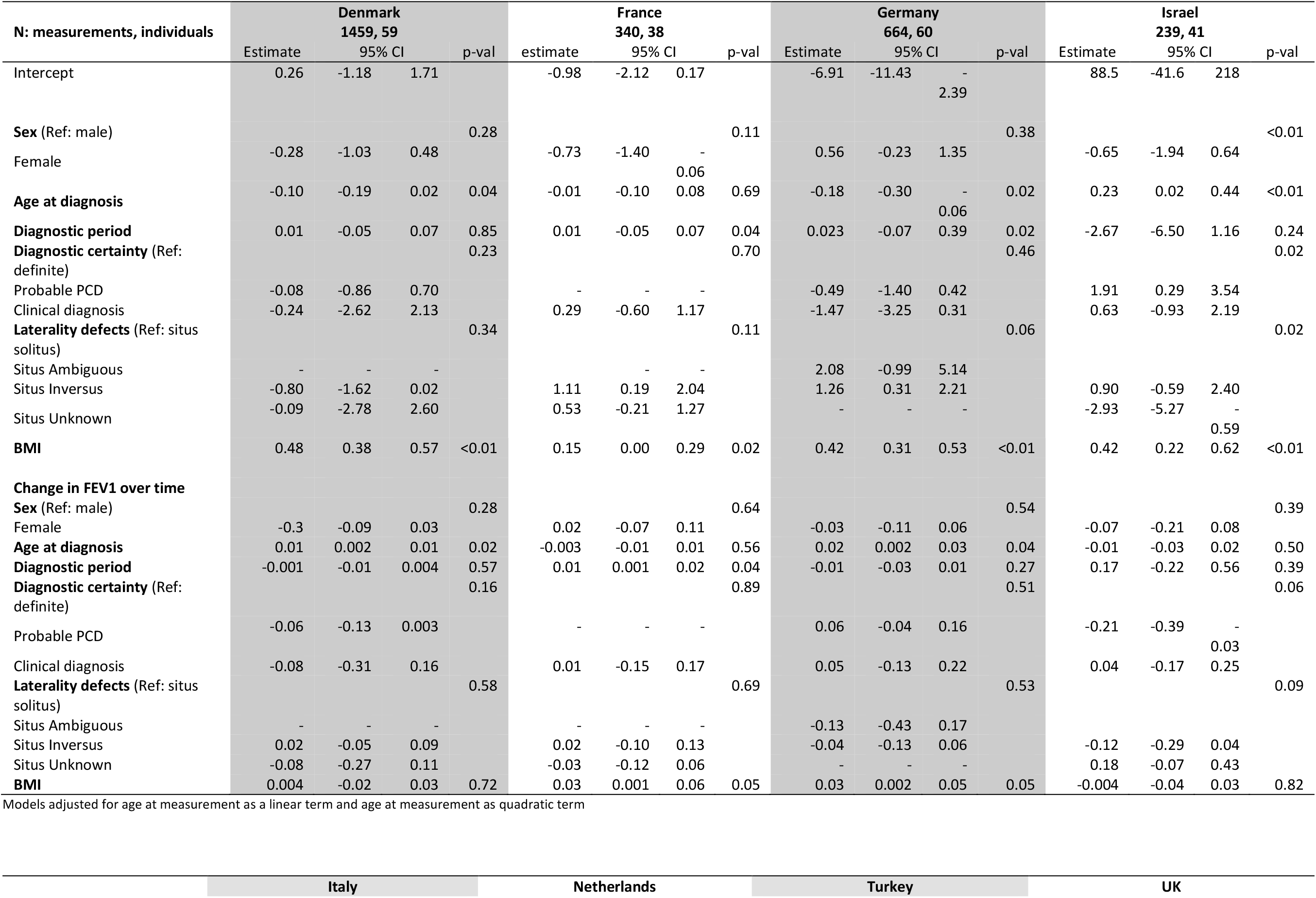
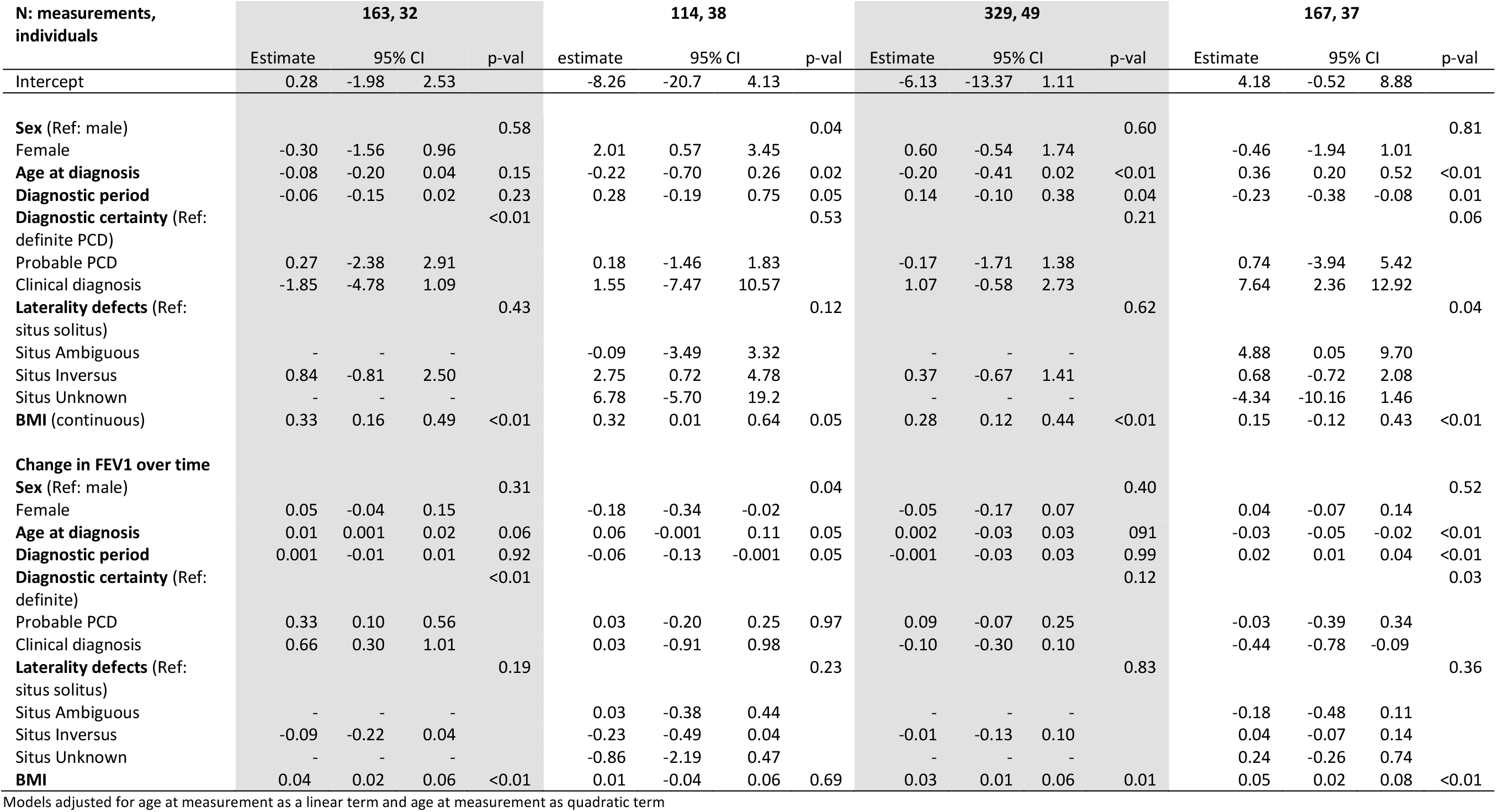
FEV1 of patients with primary ciliary dyskinesia (PCD) from the international PCD cohort compared to Global Lung Function Initiative 2012 reference values (linear mixed effects regression, adjusting for all covariates) for single countries with more than 30 patients

## Author Contributions

CE Kuehni, FS Halbeisen and M Goutaki developed the concept and designed the study. FS Halbeisen cleaned and standardised the data. FS Halbeisen and E Pedersen performed the statistical analyses. All other authors participated in discussions for the development of the study and contributed data. FS Halbeisen, CE Kuehni and M Goutaki drafted the manuscript. All authors commented and revised the manuscript. CE Kuehni and FS Halbeisen take final responsibility for the contents.

## Acknowledgments

We want to thank all the patients in the iPCD cohort and their families, and we are grateful to the PCD support groups that closely collaborate with us. We thank all the researchers in the participating centres who helped collect and enter data and worked closely with us throughout the build-up of the iPCD Cohort.

